# Detection of tau spatial extent using MK6240 and flortaucipir PET

**DOI:** 10.64898/2026.10.26.26366488

**Authors:** Guilherme Povala, Bruna Bellaver, Livia Amaral, Firoza Z. Lussier, Arthur C. Macedo, Pamela C.L. Ferreira, Douglas T. Leffa, Guilherme Bauer-Negrini, Marina Scop Medeiros, Emma Ruppert, Cécile Tissot, Thomas K. Karikari, Joseph C. Masdeu, Juan Fortea, Dana L. Tudorascu, Val Lowe, Hwamee Oh, David N. Soleimani-Meigooni, Belen Pascual, Brian A. Gordon, Pedro Rosa-Neto, Suzanne Baker, Tharick A. Pascoal, the HEAD study

## Abstract

**INTRODUCTION:** Tau PET enables in vivo quantification of neurofibrillary tangle spread, yet whether tau spatial extent (TSE) performs comparably across tracers remains unknown.

**METHODS:** We performed a head-to-head comparison of Flortaucipir and MK6240 tau PET in 682 individuals across the AD continuum, quantifying TSE as the percentage of regional voxels exceeding a tracer-specific abnormality threshold and evaluated it as a metric of tracer comparability.

**RESULTS:** MK6240 showed greater TSE than Flortaucipir (CU Aβ+, medial temporal lobe [MTL]: 42% vs 25%; dementia Aβ+, late neocortical: 55% vs 42%), reaching 50% detection probability at lower Aβ burden (Centiloid: MTL, 26 vs 45; late neocortical, 95 vs 118). TSE showed higher associations than SUVR with memory, Aβ PET, and plasma pTau217, particularly in the neo-late composite. Participants with intermediate tau involvement (TSE 25–50%), most below the SUVR positivity thresholds (MTL positivity 10.2–16.9%), had approximately three-fold higher odds of Aβ PET and pTau217 positivity than TSE 0–25% (ORs 2.90–3.46). 53.4% of intermediate Flortaucipir cases (TSE 25–50%) shifted to a higher MK6240 category, and 82.5% of moderately high Flortaucipir cases (50–75%) were classified as near-saturated (75–100%) by MK6240. Neither raw TSE nor raw SUVR was directly interchangeable across tracers, with systematic and proportional bias for both metrics (MTL Deming slope: TSE 1.29; SUVR 2.69).

**DISCUSSION:** TSE showed stronger unadjusted associations than SUVR in selected region- and outcome-specific comparisons, supporting its potential as a complementary measure. Like SUVR, raw TSE values were not directly interchangeable between flortaucipir and MK6240, and tracer-specific calibration and validation are required before cross-tracer use.

## 1. Background

The advent of tau positron emission tomography (PET) has enabled the visualization of the spatial progression of tau pathology in living individuals, with multiple studies demonstrating that in vivo regional uptake patterns recapitulate neuropathological topographic progression [1–5]. This topographic information carries important clinical significance, as the spatial distribution of tau pathology predicts future atrophy with regional specificity [6], outperforms amyloid-Aβ (Aβ) PET and magnetic resonance imaging (MRI) in predicting cognitive decline [7], and independently stratifies risk for Alzheimer’s disease (AD) progression in cognitively unimpaired (CU) individuals [8]. The revised Alzheimer’s Association diagnostic criteria formalize this principle by defining biological disease stages according to the regional tau spread, from medial temporal through moderate and widespread neocortical involvement, positioning tau topography as central to disease characterization [9].

Despite the clinical importance of tau spatial distribution, the predominant quantitative metric for tau PET, the standardized uptake value ratio (SUVR), reflects mean tracer uptake within a region of interest rather than the spatial distribution of pathology, and thus cannot distinguish focal high-intensity from diffuse moderate-intensity tau burden [10, 11]. To overcome these limitations, previous studies have recently introduced spatial extent metrics that quantify the extent of region exceeding a threshold for abnormality [10–12]. These metrics have demonstrated stronger associations with cognitive performance and neurodegeneration than SUVR [10, 13], greater sensitivity for detecting early-stage tau positivity [11], and the ability to capture biologically meaningful patterns beyond tau burden intensity alone [12]. In addition, due to differences in binding affinity, off-target binding profiles, and SUVR dynamic range, SUVR values are not directly comparable across tracers [14, 15]. Because spatial extent metrics binarize voxels with tracer-specific thresholds before computing proportions, they may in principle provide a more commensurable cross-tracer metric than intensity-based SUVR.

Although spatial extent metrics have shown promise within individual tau PET tracers, their comparability across tracers and value for tracer-aware disease characterization remain important open questions. Resolving this gap is crucial for consistent staging of tau spread and for reliable biomarker endpoints in emerging anti-tau clinical trials, where limiting further propagation may be more achievable than reversing established pathology [16]. Here, we performed a prospective, multicenter within-participant head-to-head comparison of the tau spatial extent (TSE) between Flortaucipir and MK6240 across the AD continuum, examined the relationship between TSE and tracer uptake intensity, and assessed the direct interchangeability of TSE values across tracers.

## 2. Methods

### 2.1. Study design and participants

The HEAD study (ClinicalTrials.gov ID: NCT05361382) utilized a multicenter, prospective, cross-sectional design implemented at eight research centers throughout the United States and Canada. Enrollment included young healthy volunteers (18-28 years of age) and older adults (50-90 years of age) spanning the clinical spectrum from CU through MCI to dementia. All participants provided written informed consent prior to study procedures. Each participating site obtained institutional review board (IRB) approval for the study protocol. For United States-based sites, Advarra functioned as the central IRB of record (Protocol ID: Pro00061365). Additional details regarding study design and participant characteristics are available elsewhere [15].

### 2.2. Clinical assessment and diagnostic classification

Participants completed standardized clinical and cognitive evaluations utilizing the National Alzheimer’s Coordinating Center (NACC) Uniform Data Set (UDS), either version 3.0[17] or 4.0, which encompasses the Clinical Dementia Rating (CDR) scale and an extensive neuropsychological battery including the Montreal Cognitive Assessment (MoCA). The Mini-Mental State Examination (MMSE) was also performed for all enrollees. CU older adults were aged 50 years or above with CDR global scores of 0 [18]. Site investigators documented the presence of cognitive impairment through clinical evaluation. The MCI diagnostic category required both subjective reports and objective demonstration of cognitive decline, CDR scores of 0.5, and maintenance of functional independence in most activities of daily living. The mild to moderate dementia group satisfied diagnostic criteria established by the National Institute on Aging-Alzheimer’s Association [18] and demonstrated CDR scores ranging from 0.5 to 2. A memory composite score was computed as the mean of standardized z-scores from domain-specific neuropsychological tests, normed against a reference sample of CU individuals with normal global cognition (CDR = 0, MoCA ≥ 27, MMSE ≥ 26) [19]. The memory composite comprised immediate and delayed recall scores from the Craft Story 21 Recall and the Benson Complex Figure tests.

### 2.3. MRI and PET image acquisition and processing

MRI scanning utilized 3T systems with harmonized protocols according to ADNI4 standards [20]. High-resolution T1-weighted images were acquired using 3D MPRAGE or IRSPGR sequences. All participants completed head-to-head Aβ and two tau PET imaging. Aβ tracers included [^11^C]PiB, [^18^F]Florbetaben, or [^18^F]NAV4694. Tau imaging employed [^18^F]MK6240 and [^18^F]Flortaucipir in head-to-head acquisitions (mean time between scans = 33 ± 44 days). For each participant, flortaucipir and MK6240 PET images were acquired at the same imaging site using the same scanner. This within-participant, head-to-head acquisition design ensured that scanner- and site-specific factors were held constant for comparisons between tracers and for comparisons of TSE with regional SUVR. Acquisition windows were: PiB (40–60 or 50–70 min), Florbetaben (90–110 min), NAV4694 (50–70 min), MK6240 (90–110 min), and Flortaucipir (80–100 min) post-injection. Images were reconstructed into four 5-minute frames using OSEM-based algorithms (VPHD, VPFX, TOF-OSEM) with standardized parameters and no smoothing, and standard corrections (attenuation, motion, dead time, decay, scatter, random events) were applied. Scanner models, reconstruction, and correction parameters are available in **Supplemental Table 1**.

PET images were processed using our previously described and validated MRI-based pipeline [21], comprising T1-weighted preprocessing and segmentation, cerebellar parcellation, PET-to-MRI co-registration, nonlinear normalization to ICBM 2020 Nonlinear Symmetric space [22], and visual quality control of all registrations. Reference regions were the native-space inferior cerebellar grey matter for tau PET and the whole cerebellum for Aβ PET, as recommended for Centiloid calibration [23]. Scanner-specific Gaussian smoothing was applied to achieve a final 8 mm FWHM effective resolution (**Supplemental Methods)**. Regional SUVRs were extracted as median values [24] within CerebrA atlas ROIs [25], modified to incorporate a validated transentorhinal segmentation in 1-mm isotropic space [26, 27]. Centiloid calculation employed the standard composite ROI (frontal, temporal, parietal cortices, precuneus, anterior striatum, insula) [23]. Individuals were considered Aβ PET positive if had Centiloid values higher than 20 or positive on visual reads.

TSE was derived to quantify the spatial extent of tau pathology. Voxel-wise thresholds for abnormality were established using the young control cohort (aged 18-28 years) as a reference population, defined as the mean SUVR plus 2.5 standard deviations (SD) for each tracer [28]. Binary abnormality maps were generated for each participant by classifying voxels exceeding this threshold as abnormal. TSE was calculated as the proportion of abnormal voxels within each ROI relative to the total number of voxels in that region, expressed as a percentage, ranging from 0 to 100%. As a sensitivity analysis, TSE was recomputed using Youden threshold and young-reference mean + 2 SD, + 2.5 SD and + 3 SD thresholds. TSE region positivity was defined as ≥25% of voxels within the region exceeding the abnormality threshold, unless otherwise specified. As used for TSE, positivity thresholds for SUVR and plasma biomarkers were defined as the young reference group mean plus 2.5 SD, after removing outliers beyond 3 SD [15].

Modified Braak-like staging utilized hierarchical ROI groupings: Braak I (transentorhinal cortex), II (entorhinal cortex and amygdala), III (parahippocampal, fusiform, and lingual gyri), IV (inferior and middle temporal cortices, caudal and rostral anterior cingulate, isthmus and posterior cingulate, and insula), V (frontal, parietal, and occipital cortices, transverse and superior temporal gyri, precuneus; pars opercularis, triangularis and orbitalis, supramarginal gyrus), and VI (precentral, postcentral, and paracentral gyri, cuneus; and pericalcarine cortex) [2]. Composite ROIs included the MTL (entorhinal cortex and amygdala) [8] and neo-late (Braak V-VI combined, closely corresponding to the FDA-approved read region for Flortaucipir).

### 2.4. Plasma pTau217 quantification

Plasma collection and tau-PET acquisition were performed within 45 days of each other. Blood was drawn into EDTA tubes following NRCAD guidelines, and plasma was aliquoted and stored at −80 °C. Samples were shipped under temperature-controlled conditions to the University of Pittsburgh and thawed once prior to analysis. Plasma pTau217 concentrations were measured using the ALZpath Advantage PLUS assay on the single-molecule array (Simoa) platform (Quanterix).

### 2.5. Statistical analyses

Between-tracer TSE differences across clinical, Aβ and cognitive severity strata were tested with paired Wilcoxon signed-rank tests. Differences in TSE positivity rates were tested with McNemar’s test. Cumulative tau-positivity curves with prevalence ratios were quantified from modified Poisson regression (log link, HC0 robust standard errors). Logistic regression modeled the probability of tau positivity as a function of Centiloid. To compare detection between tracers, we calculated the NND as the reciprocal of the paired risk difference (proportion positive on MK6240 but negative on Flortaucipir, minus the reverse). Confidence intervals were estimated using 1,000 bootstrap resamples, with significance defined as a 95% CI for the risk difference excluding zero. The SUVR–TSE relationship was assessed with Spearman correlations, and tracer-specific dynamic range was evaluated among participants with regional spatial saturation (TSE ≥ 75%). To compare TSE and SUVR for early detection, Spearman correlations with clinical and biomarker outcomes were compared using the Steiger z-test applied to ranked data, and Cohen’s d effect sizes for group separations were compared after d-to-r conversion, with bootstrap 95% confidence intervals (1,000 resamples) were computed for all estimates. To evaluate AD-related biomarker associations among participants with intermediate TSE who were negative according to the regional SUVR criterion, logistic regression modeled the probability of SUVR positivity as a function of TSE, biomarker levels were compared between the 0–25% and 25–50% TSE bins with Wilcoxon rank-sum tests, and odds ratios for Aβ and pTau217 positivity were estimated with Wald confidence intervals. Cross-tracer association was assessed using Spearman correlation; this analysis was interpreted as reflecting rank-order association and not numerical agreement or direct interchangeability. Cross-tracer agreement between tracers was further evaluated at the participant level using Bland–Altman analyses, including the mean paired difference (MK6240 minus flortaucipir) and 95% limits of agreement (mean difference ±1.96 SD of paired differences). Proportional bias was assessed by linear regression of paired differences on paired means. We additionally estimated Lin’s concordance correlation coefficient (CCC) and performed Deming regression, assuming equal measurement-error variance between tracers. Departure of the Deming intercept and slope from 0 and 1, respectively, was interpreted as evidence against direct interchangeability of uncalibrated measurements. Confidence intervals were obtained from 2,000 participant-level bootstrap resamples. Normalized root mean square error (nRMSE) was calculated separately for each metric and region as:

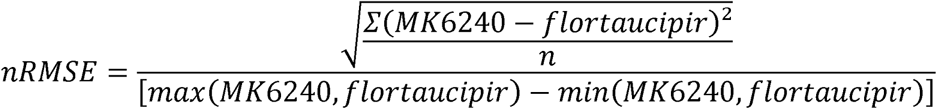

where *i* indexes participants with paired scans, n is the number of paired observations, and the denominator is the pooled observed range across both tracers. Lower nRMSE indicates lower normalized raw cross-tracer discrepancy within the analyzed sample. Cross-classification into prespecified TSE categories was also evaluated. Two-sided p < 0.05 was considered significant. Analyses were performed in R 4.4.0.

## 3. Results

### 3.1. Participants

Between March 2022 and August 2025, 775 participants were recruited across eight North American sites for head-to-head Flortaucipir and MK6240 imaging. After 70 withdrawals, 22 incomplete assessments, and 1 quality-control exclusion, 682 participants were included: 38 young adults, 362 CU older adults (21% Aβ+), 183 with mild cognitive impairment (MCI; 59% Aβ+), and 99 with dementia (88% Aβ+). Demographic and clinical characteristics of participants are summarized in **Supplemental Table 2**.

### 3.2. Tau expands stereotypically but differs between tracers

TSE was calculated as the proportion of voxels within a brain region exceeding a tracer-specific abnormality threshold (mean SUVR + 2.5 SD of young individuals), expressed as a percentage from 0 to 100%. For both tracers, regional tau involvement followed a stereotyped medial temporal–to–neocortical gradient across clinical stages, consistent with Braak staging (**Fig. 1; Supplemental Fig. 1**). However, MK6240 showed greater regional involvement than Flortaucipir across stages (**Fig. 1**; **Supplemental Table 3**). Differences were evident early in the continuum, with CU Aβ+ individuals showing higher mean MK6240 TSE in Braak I and II than Flortaucipir (49% vs 31% and 41% vs 25%, respectively; both p < 0.001; **Fig. 1b**; **Supplemental Table 3**). This divergence widened with disease progression, with MK6240 exceeding Flortaucipir across all six Braak regions in MCI and dementia Aβ+ groups, from early regions (*e.g*., Braak I in MCI: 82% vs 65%) to late neocortical regions (*e.g*., Braak VI in dementia: 44% vs 26%; all p < 0.001; **Fig. 1b**; **Supplemental Table 3**).

**Figure 1.**
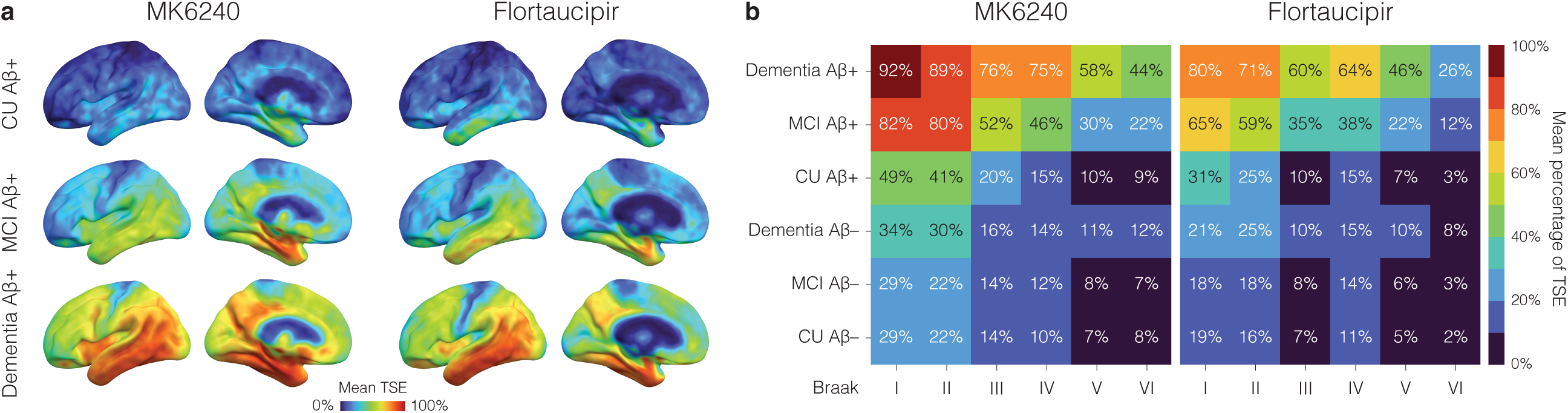
Topographical distribution of abnormal tau spatial extent for MK6240 and Flortaucipir follows similar stereotypical patterns. **(a)** Cortical surface maps display mean tau spatial extent (TSE) for MK6240 and Flortaucipir across amyloid-β positive (Aβ+) clinical groups (cognitively unimpaired [CU], mild cognitive impairment [MCI], dementia). **(b)** Heatmaps show mean TSE across Braak regions (I–VI) stratified by clinical diagnosis and Aβ status for both tracers.

### 3.3. MK6240 detects greater tau spatial extent across disease severity

Among Aβ+ individuals, MK6240 showed greater medial temporal lobe (MTL) TSE than Flortaucipir across all clinical groups (CU: 42% vs 25%; MCI: 80% vs 59%; dementia: 89% vs 72%; all p < 0.001; **Fig. 2a**; **Supplemental Table 4**). This emerged early in Aβ accumulation, exceeding Flortaucipir at low Centiloid values and widening at 10–30 (32% vs 23%, p = 0.028), 30–50 (47% vs 28%), 50–75 (68% vs 43%), and >100 (91% vs 72%; all p < 0.001; **Fig. 2b**; **Supplemental Table 5**). A similar pattern was seen across cognitive severity, from clinical dementia rating sum of boxes (CDR-SB) 0 (21% vs 14%) to 0.5–1 (47% vs 35%) and ≥4.5 (82% vs 67%; all p < 0.001; **Fig. 2c**; **Supplemental Table 6**). Accordingly, more participants exceeded every MTL TSE threshold with MK6240 than Flortaucipir (≥25% TSE: 337 vs 280; ≥50% TSE: 248 vs 162; ≥75% TSE: 197 vs 105; all McNemar p < 0.001; **Fig. 2d**; **Supplemental Table 7**). The same tracer-dependent pattern was observed in the late neocortical (neo-late) composite, albeit with lower overall TSE values. MK6240 showed greater neo-late TSE across all Aβ+ groups (CU: 10% vs 7%; MCI: 28% vs 20%; dementia: 55% vs 42%; all p < 0.001; **Fig. 2e**; **Supplemental Table 4**), exceeded Flortaucipir most consistently from Centiloid 50–75 onward (**Fig. 2f**; **Supplemental Table 5**), and remained higher across CDR-SB strata (**Fig. 2g**; **Supplemental Table 6**). More participants also exceeded every neo-late threshold with MK6240 (≥25% TSE: 150 vs 110; ≥50% TSE: 72 vs 48; ≥75% TSE: 38 vs 14; all McNemar p < 0.001; **Fig. 2h**; **Supplemental Table 7**). Post-hoc sex-stratified analyses showed the same pattern, with higher MK6240 than flortaucipir TSE in both male and female participants in the MTL and neo-late composite (all within-sex paired p < 0.001), with no evidence that the tracer difference differed by sex (MTL: p = 0.400; neo-late: p = 0.973; **Supplemental Table 8**). In sensitivity analyses using alternative voxelwise thresholds, group ordering and the direction and significance of MK6240–flortaucipir differences were preserved in both regions (**Supplemental Fig. 2**).

**Figure 2.**
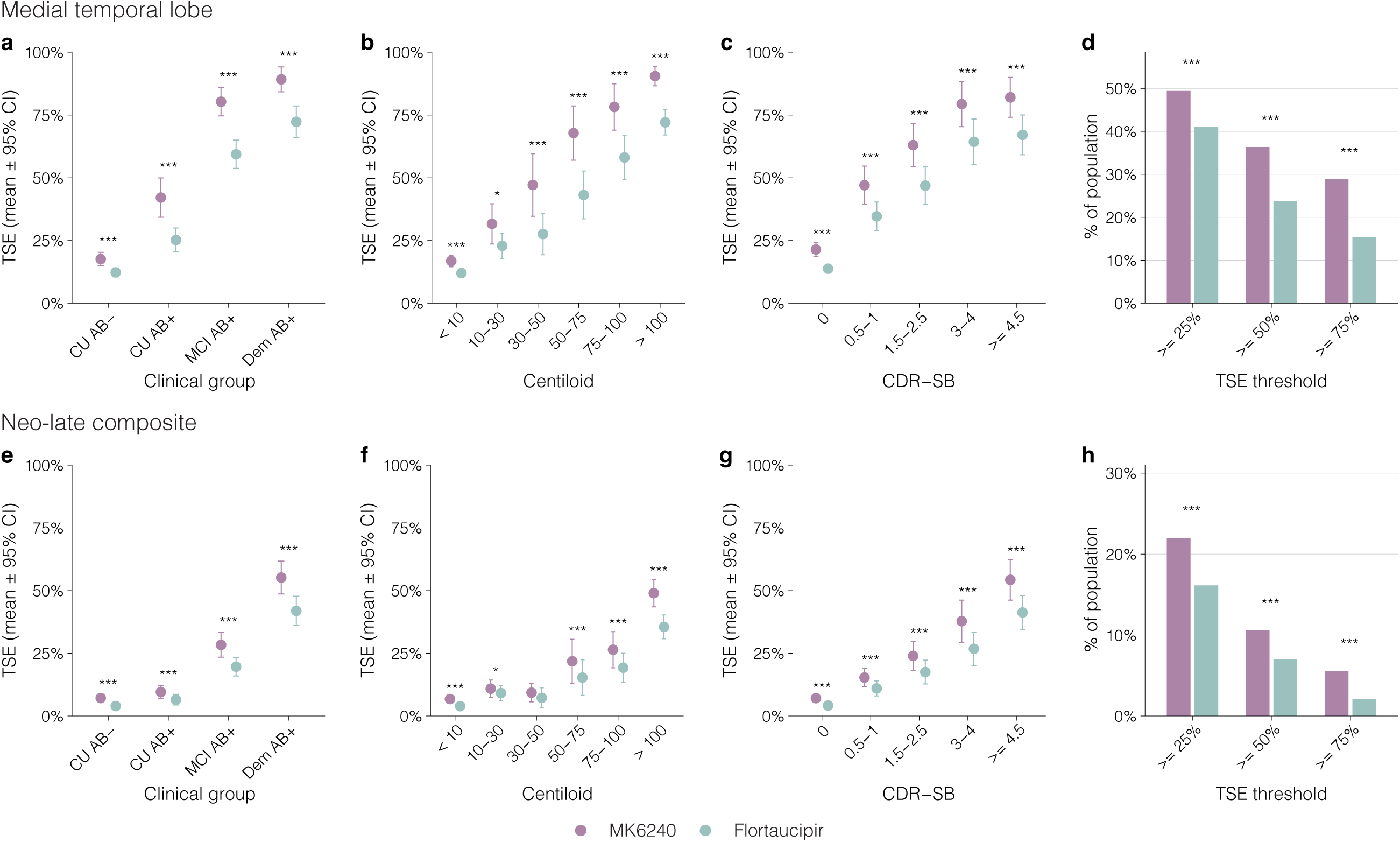
Tau PET spatial extent changes as a function of clinical severity and amyloid-β burden. Mean tau spatial extent (TSE; ±95% CI) in (**a–d**) the medial temporal lobe (MTL) and (**e–h**) neo-late composite across (**a,e**) clinical groups, (**b,f**) Centiloid bins, and (**c,g**) CDR-SB bins. (**d,h**) Bar plots show the percentage of the population exceeding TSE thresholds of 25%, 50%, and 75% for each tracer. Asterisks denote between-tracer differences (paired Wilcoxon signed-rank test for **a–c,e–g**; McNemar test for **d,h**). * *P* < 0.05; ** *P* < 0.01; *** *P* < 0.001. CDR-SB: Clinical Dementia Rating Sum of the Boxes; CU: cognitively unimpaired; MCI: mild cognitive impairment.

### 3.4. Prevalence of tau positivity was higher with MK6240 than Flortaucipir

Cumulative tau-positivity curves using modified Poisson regression with robust standard errors confirmed higher tau-positivity prevalence (TSE ≥25%) with MK6240 than Flortaucipir in CU participants in the MTL (prevalence ratio [PR] = 1.42, p = 0.026) and in cognitively impaired participants in the MTL (PR = 1.11, 95% CI 1.04–1.19, p = 0.002) and neo-late composite (PR = 1.23, 95% CI 1.04–1.46, p = 0.015; **Fig. 3a**; **Supplemental Table 9**). The lack of a significant difference in cognitively unimpaired participants in the neo-late composite likely reflected the low prevalence of neo-late TSE positivity in this subgroup. MK6240 reached 50% detection probability at lower Aβ burden than Flortaucipir (Centiloid: MTL, 26 vs 45; neo-late, 95 vs 118; **Fig. 3b**; **Supplemental Table 10**). Accordingly, the number of individuals needed to detect one tau-positive case was consistently lower with MK6240 across clinical groups (CU Aβ+: MTL, 1.8 vs 2.4; neo-late, 8.8 vs 22.0; MCI Aβ+: 1.1 vs 1.2 and 2.3 vs 3.1; dementia Aβ+: 1.0 vs 1.1 and 1.3 vs 1.5; **Fig. 3c**). Bootstrapped number needed to detect (NND) analyses showed that using MK6240 instead of Flortaucipir identified one additional tau-positive case for every 6.9, 9.8, and 12.2 individuals screened in the MTL (CU Aβ+, MCI Aβ+, and dementia Aβ+, respectively), and every 9.0 and 8.8 screened in the neo-late composite (MCI Aβ+ and dementia Aβ+; all p < 0.05; **Fig. 3d**; **Supplemental Table 11**).

**Figure 3.**
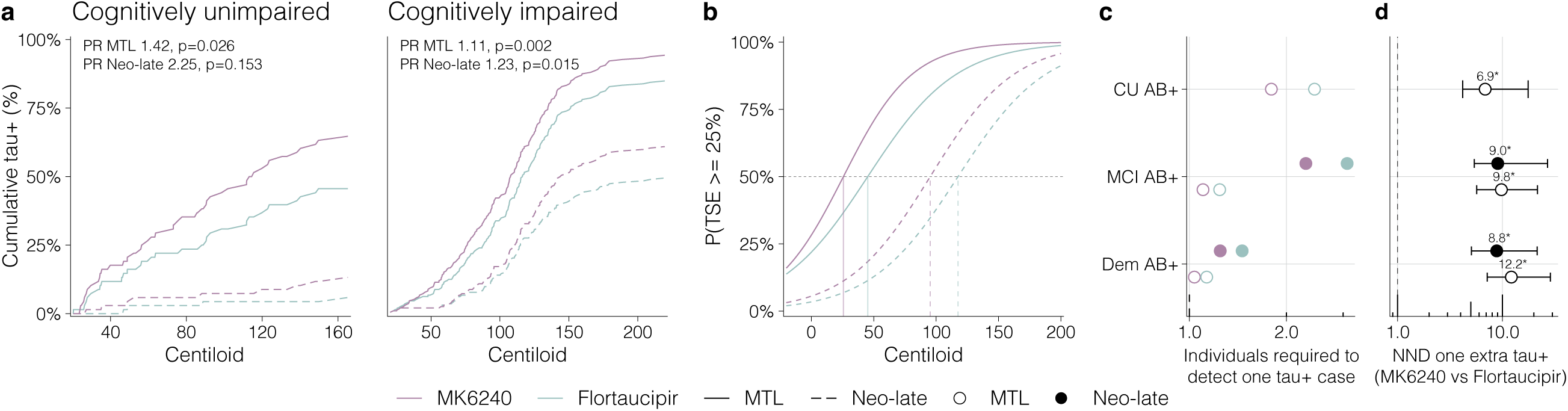
Comparison of TSE positivity prevalence and detection thresholds for MK6240 versus Flortaucipir across. **A**β **burden. (a)** Cumulative tau-positivity curves as a function of Centiloid for Aβ+ cognitively unimpaired (CU) and cognitively impaired (mild cognitive impairment [MCI] and dementia together) groups, shown separately for medial temporal lobe (MTL; solid lines) and neo-late composite (dashed lines). Prevalence ratios (PR) with robust standard errors from modified Poisson regression are displayed. **(b)** Logistic regression curves modeling the probability of tau spatial extent (TSE) ≥ 25% as a function of Centiloid for each tracer–region combination; vertical lines indicate the Centiloid at which P = 50%. **(c)** Number of individuals required to detect one tau-positive case, by clinical group, tracer and brain region. Open circles denote MTL; filled circles denote neo-late. **(d)** Bootstrapped number needed to detect (NND) one additional tau-positive case with MK6240 versus Flortaucipir, with 95% confidence intervals.

### 3.5. TSE provides complementary information to SUVR across tracers

SUVR and TSE were strongly correlated across regions for both tracers (Spearman Rho = 0.81–0.97; all p < 0.0001; **Fig. 4a–d**), with TSE rising steeply at low SUVR values before plateauing near 100%. Specifically, at high MTL involvement (TSE ≥75%), MK6240 SUVR continues to increase, retaining a broad dynamic range and consequently greater data dispersion (IQR 1.056; min 1.08; max 4.51), whereas Flortaucipir SUVR increases are more modest (IQR 0.240; min 1.42; max 2.33; **Supplemental Table 12**). At a lower tau burden, individuals classified as negative according to the regional SUVR positivity thresholds showed TSE values ranging from 0% to greater than 50%, indicating that intermediate TSE was present despite SUVR negativity (**Fig. 4e–h**). TSE showed stronger associations than SUVR with memory, Centiloid, and plasma pTau217, particularly in the neo-late composite for both tracers across these outcomes (all p < 0.001; **Fig. 4i, j**; **Supplemental Table 13**). TSE also showed larger effect sizes than SUVR for selected clinical-group contrasts (**Fig. 4k-l**). In the MTL, TSE exceeded SUVR for contrasts such as MCI Aβ+ versus CU Aβ− and cognitively impaired Aβ− for both tracers (**Fig. 4k**; **Supplemental Table 14**). In the neo-late composite, TSE yielded significantly larger effect sizes for the examined cognitively impaired group comparisons in the neo-late composite, reaching up to 1.8-fold greater separation than SUVR (**Fig. 4l**; **Supplemental Table 14**).

**Figure 4.**
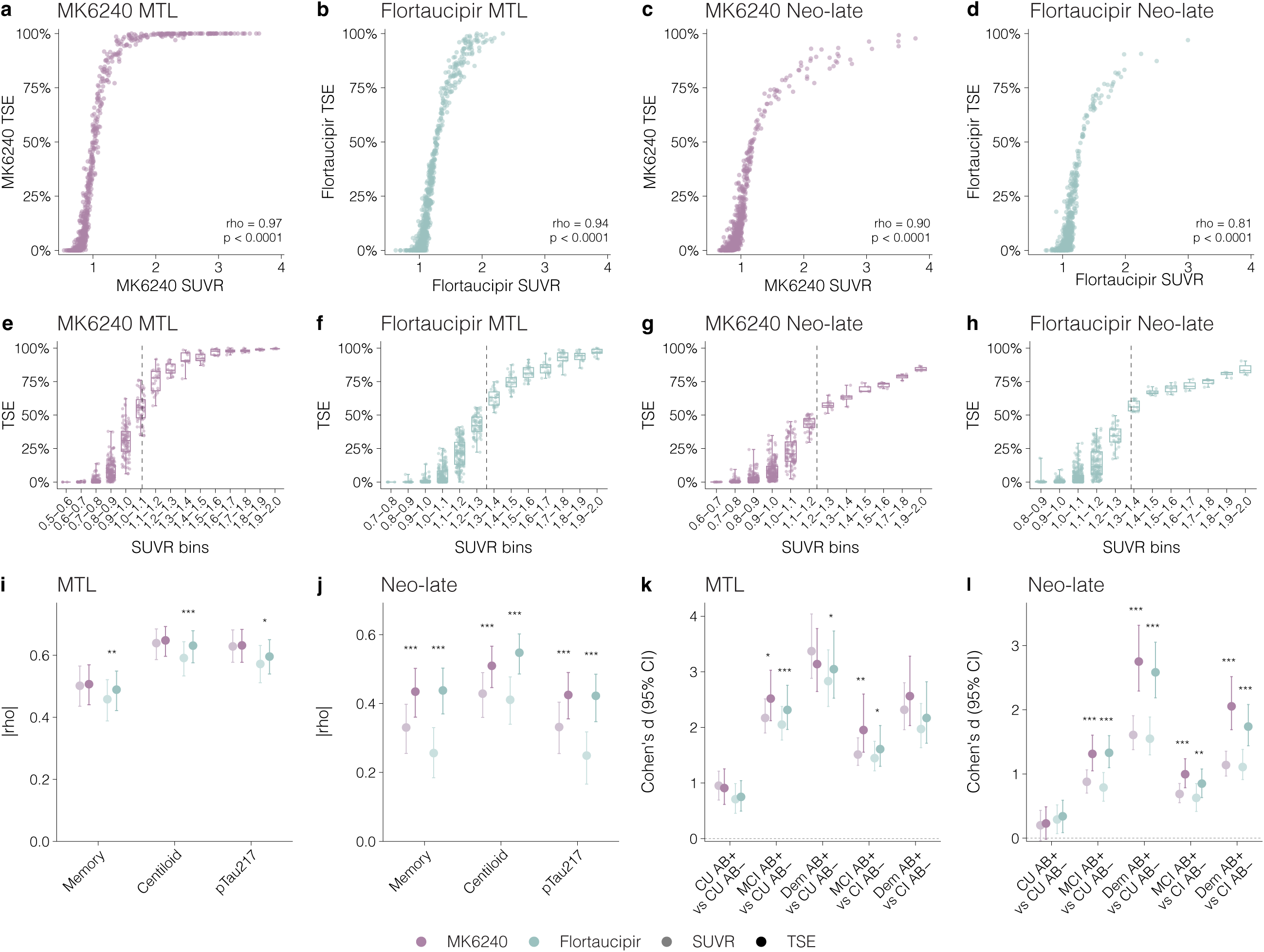
Comparison of TSE and SUVR for MK6240 and Flortaucipir quantification. **(a–d)** Scatterplots show the relationship between standardized uptake value ratio (SUVR) and tau spatial extent (TSE) for MK6240 and Flortaucipir in medial temporal lobe (MTL) and neo-late composite regions, with points colored by Aβ burden (Centiloid). Spearman Rho and *P* values are shown. **(e–h)** Box plots display TSE distributions within SUVR bins; dashed vertical lines indicate SUVR positivity thresholds. **(i,j)** Forest plots compare absolute Spearman correlations (|Rho|) of TSE versus SUVR with memory composite, Centiloid and plasma pTau-217 for each tracer in the MTL and neo-late regions. Bootstrap 95% CIs are shown. **(k,l)** Forest plots compare Cohen’s *d* effect sizes (with bootstrap 95% CIs) for clinical group separations between TSE and SUVR. Asterisks denote significant TSE-SUVR differences (Steiger *z*-test). * *P* < 0.05; ** *P* < 0.01; *** *P* < 0.001. CU: cognitively unimpaired; MCI: mild cognitive impairment.

### 3.6. TSE is associated with AD-related biomarkers below SUVR positivity thresholds across tracers

Logistic models showed that the threshold for 50% SUVR positivity corresponded to similar MTL TSE levels for both tracers (MK6240: 49%; Flortaucipir: 47%; **Fig. 5a**). SUVR positivity rates remained near zero in the 0–25% TSE bin, with a small percentage SUVR-positive at 25–50% TSE (MK6240:16.9%, Flortaucipir: 10.2%), and virtually all individuals SUVR-positive above 50% TSE (**Fig. 5b**; **Supplemental Table 15**). Participants in this early TSE range (25–50%) already showed significantly elevated Centiloid and plasma pTau217 versus the 0–25% bin for both tracers (all p < 0.01; **Fig. 5c**, **d**; **Supplemental Table 16**), with significantly higher odds of Aβ positivity (MK6240: OR = 3.46, 95% CI 1.94–6.18; Flortaucipir: OR = 2.92, 95% CI 1.85–4.62; both p < 0.001) and pTau217 positivity (MK6240: OR = 2.90, 95% CI 1.68–5.00, p < 0.001; Flortaucipir: OR = 3.02, 95% CI 1.94–4.71, p < 0.001; **Fig. 5e**). Results were consistent when restricting to SUVR-negative individuals (**Supplemental Table 17**).

**Figure 5.**
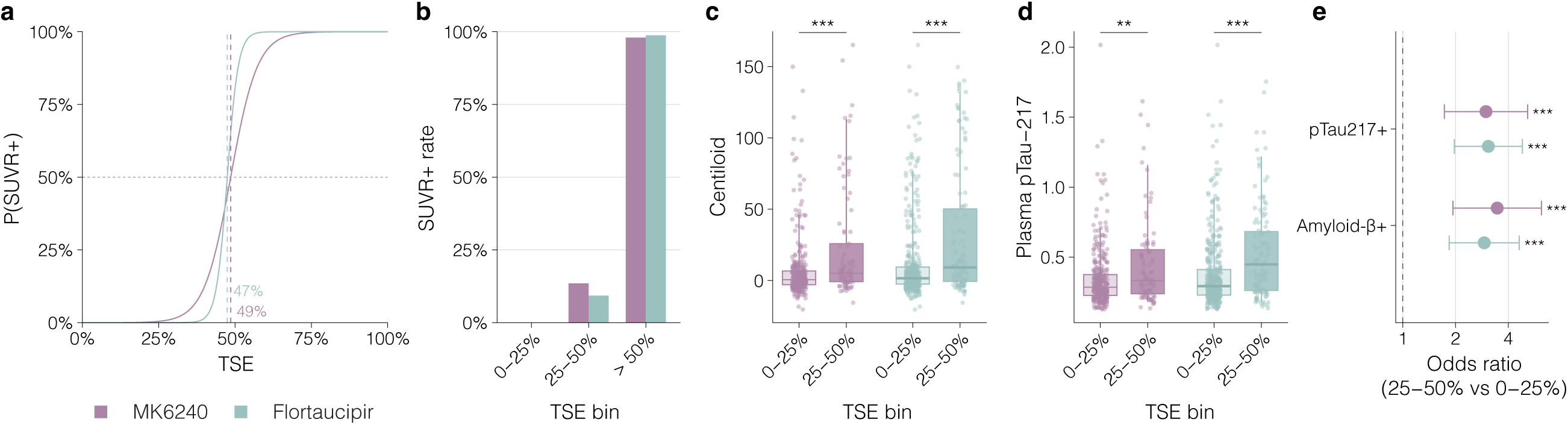
TSE is associated with AD-related biomarkers below conventional SUVR positivity thresholds. (**a**) Logistic regression curves modelling the probability of SUVR positivity as a function of MTL TSE for each tracer; dashed vertical lines indicate the TSE value at which P(SUVR+) = 50%, which was nearly identical for both tracers. (**b**) SUVR positivity rates within TSE bin showed that positivity rates remain near zero in the 0–25% TSE range, rise modestly in the 25–50% range, and approach universality above 50% TSE. (**c, d**) Box plots compare Centiloid (**c**) and plasma pTau-217 (**d**) between participants in the 0–25% and 25–50% TSE bins, separately for each tracer (Wilcoxon rank-sum test). (**e**) Forest plot showing odds ratios (95% CI) for amyloid-β (Aβ) positivity and pTau-217 positivity in the 25–50% TSE bin relative to the 0–25% TSE bin, estimated from logistic regression models. Individuals in the 25–50% TSE range have significantly higher odds of Aβ positivity and plasma pTau-217 positivity relative to those in the 0–25% range. * *P* < 0.05; ** *P* < 0.01; *** *P* < 0.001.

### 3.7. TSE values are not directly interchangeable across tau PET tracers

Participant-level TSE correspondence between tracers was high (MTL: Rho = 0.877; neo-late: Rho = 0.789; p < 0.0001) but this association did not imply numerical agreement or direct interchangeability (**Fig. 6a, e; Supplemental Table 18**). In the MTL, upward reclassification by MK6240 was evident even at minimal Flortaucipir involvement (0–25%: 20.6% reclassified higher), increased at intermediate levels (25–50%: 53.4% higher, 25.4% same, 21.2% lower), and was near-universal at moderately high Flortaucipir involvement (50–75%: 82.5% reclassified into the highest MK6240 bin; **Fig. 6b, c; Supplemental Table 19**). All participants at maximum Flortaucipir involvement (75–100%) were also classified as maximal by MK6240; however, when conditioning on MK6240 TSE of 75–100%, only 53.3% were classified in the same category by flortaucipir, whereas 46.7% were classified lower (**Fig. 6d; Supplemental Table 20**). Although ceiling effects were less pronounced for MK6240 TSE when examined as a function of flortaucipir TSE, the same directional pattern was observed in the neo-late composite (0–25%: 9.1% reclassified higher; 25–50%: 35.5% higher, 48.4% unchanged, 16.1% lower; 50–75%: 67.6% reclassified as maximal; among participants with MK6240 TSE of 75–100%, only 36.8% were classified in the same category by flortaucipir and 63.2% were classified lower; **Fig. 6f; Supplemental Table 20**). Paired method-comparison analyses further demonstrated systematic and magnitude-dependent cross-tracer differences for both TSE and SUVR. For TSE, the mean MK6240–flortaucipir difference was 10.6% in the MTL (95% CI, 9.1% to 12.0% **Fig. 6g**) and 5.0% in the neo-late composite (95% CI, 4.3% to 5.9%, **Fig. 6h**), with proportional bias in both regions (both p < 0.001, **Supplemental Table 21**). Deming regression slopes differed from 1 for TSE in the MTL (1.285; 95% CI, 1.241 to 1.336) and neo-late composite (1.295; 95% CI, 1.244 to 1.361, **Supplemental Table 22**). SUVR similarly showed marked proportional bias and Deming slopes inconsistent with identity in the MTL (2.686; 95% CI, 2.536 to 2.839) and neo-late composite (2.673; 95% CI, 2.445 to 2.931, **Supplemental Table 22**). nRMSE was higher for TSE than SUVR in the MTL (21.8% [95% CI, 20.2% to 23.3%] versus 10.8% [95% CI, 9.8% to 11.9%]) and neo-late composite (11.7% [95% CI, 10.8% to 12.8%] versus 7.0% [95% CI, 5.7% to 8.2%], **Supplemental Table 23**). Conversely, CCC was higher for TSE than SUVR (MTL: 0.824 vs 0.628; neo-late: 0.858 vs 0.619; **Supplemental Table 22**).

**Figure 6.**
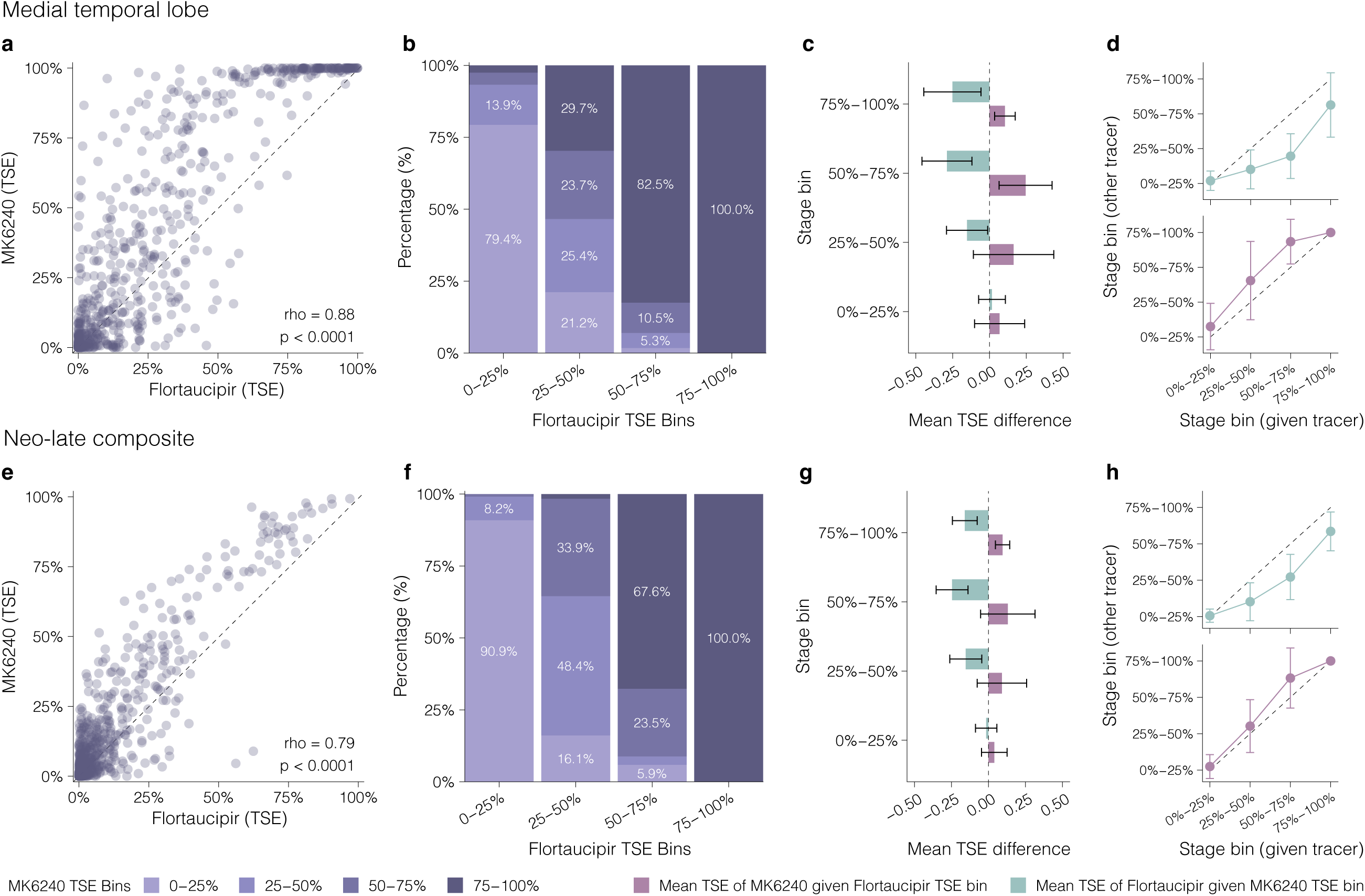
TSE is correlated across tracers, but not interchangeable. **(a,e)** Scatterplots show participant-level correspondence of tau spatial extent (TSE) between tracers in the medial temporal lobe (MTL) and neo-late composite, with Spearman Rho and *P* values. **(b,f)** Stacked bar charts display cross-classification of MK6240 TSE bins within each Flortaucipir TSE bin, with segment percentages shown. **(c,g)** Bar plots show mean TSE difference (± SD) between tracers within each TSE stage bin, conditioned separately on MK6240 bins (mean Flortaucipir TSE given MK6240 bin) and Flortaucipir bins (mean MK6240 TSE given Flortaucipir bin). The dashed line indicates zero difference. **(d,h)** Mean TSE (± SD) stage of the other tracer as a function of the given tracer’s TSE stage bin, shown separately for each conditioning direction.

## 4. Discussion

In this large multicenter, within-participant head-to-head study across the AD continuum, we show that tau spatial extent is clinically informative but not tracer invariant. TSE showed stronger associations with memory, Aβ burden, and plasma pTau217 in selected comparisons, and showed larger effect sizes than SUVR for selected clinical-stage contrasts. MK6240 consistently showed broader MTL and neocortical involvement than Flortaucipir at comparable levels of Aβ pathology and clinical severity. This systematic offset produced substantial cross-tracer discordance, most pronounced at intermediate stages of tau pathology where staging and treatment decisions may be most consequential. Critically, neither raw TSE nor raw SUVR was directly interchangeable across tracers. Both showed systematic and magnitude-dependent bias, indicating that tracer-specific calibration is required before either metric is compared across tracers.

We observed that MK6240 showed greater TSE across brain regions than Flortaucipir. Although competitive binding assays have demonstrated overlapping binding sites on paired helical filament tau for both tracers, postmortem studies reporting higher total binding of MK6240 to AD brain tissue in tangle-bearing cortical regions, together with its higher affinity for tau aggregates than Flortaucipir, may partly account for the broader spatial extent observed here [29, 30]. However, overlapping binding profiles do not necessarily result in identical in-vivo signal properties. Differences in binding-site accessibility, regional kinetics, dynamic range, and off-target signal may influence voxelwise threshold classification, particularly in the MTL. Flortaucipir off-target binding in the choroid plexus and the distinct signal properties of MK6240 may further influence voxelwise classification near medial temporal structures. The present data cannot determine the relative contribution of these factors, nor do they establish preferential MK6240 binding to pretangles or other distinct tau species, which will require neuropathological or autoradiographic validation. This distinction has direct implications for clinical implementation as the same patient may show 25–50% regional involvement with one tracer yet near-maximal involvement (≥75%) with another, leading to different disease stage assignments based on this metric. If anti-amyloid therapies are most effective before tau PET has spread widely across the cortex [31], Flortaucipir may classify some individuals as having residual potential for future spread, whereas MK6240 indicates that neocortical involvement is already extensive.

Region-specific TSE showed stronger associations than SUVR with memory impairment, Aβ PET burden, and plasma pTau217 in the neo-late composite for both tracers. In the MTL, however, the relative advantage of spatial spread quantification was tracer-specific, with TSE showing stronger associations than SUVR for Flortaucipir but not for MK6240. This dissociation likely reflects differences in tracer binding characteristics. For example, the already high MK6240 uptake signal within the relatively small MTL region may render average voxel intensity a sufficiently robust measure in this region. In larger neocortical regions, where intensity-based measures average signal across areas only partially infiltrated by tau pathology, both tracers may benefit from spatial extent quantification, as TSE may be sensitive to focal abnormal signal that is diluted by regional averaging and remains below the SUVR positivity threshold. The earlier involvement and potential ceiling effects in the MTL may further reduce interindividual variability and attenuate associations with clinical and biomarker measures. Consistent with this interpretation, a recent multi-cohort study demonstrated that TSE detects longitudinal tau changes in regions where SUVR changes are attenuated or absent, supporting the idea that spatial extent captures sub-threshold focal signal [32]. Recent work in two independent cohorts reported that tau PET fill state provided greater diagnostic discrimination of cognitive impairment than SUVR [33], supporting spatial extent as a potentially complementary measure of tau pathology. In the present study, however, uncalibrated TSE was not directly interchangeable between flortaucipir and MK6240, indicating that the clinical utility and cross-tracer comparability of spatial-extent measures require separate validation. Consequently, the optimal contexts of use for TSE may differ between tracers, and tracer-specific validation of spatial extent thresholds will be an important consideration before these metrics can be broadly implemented in clinical or trial settings.

These findings have direct implications for the implementation of the Alzheimer’s Association revised diagnostic criteria [9], which define biological stages based on the topographic extent/spread of tau pathology. The systematic discordance between tracers suggests that individuals could be classified into different stages depending on which tracer is used. At intermediate TSE levels in the MTL (25–50%), about 25% of participants were assigned to the same category by both tracers. Given the substantial variability across tau PET tracers, the implementation of spatial extent metrics in clinical settings will require careful standardization. Before such measures can be reliably applied in a multi-tracer environment, whether for disease staging or treatment eligibility, tracer-specific thresholds or robust cross-tracer calibration procedures will need to be established and validated.

Whether spatial extent metrics can support cross-tracer comparison is a question of growing discussion and practical importance as multiple tau PET tracers enter research and clinical use. TSE, by binarizing voxels at tracer-appropriate thresholds before computing regional proportions, could in principle reduce some effects of tracer-specific binding properties. Our results instead show that raw TSE values are not directly interchangeable across tracers. Range-normalized cross-tracer discrepancy was higher for TSE than for SUVR (nRMSE ratio 2.02 in the MTL and 1.69 in the neo-late composite), whereas SUVR showed larger proportional differences between tracers, reflected in its lower CCC and steeper Deming slopes. Cross-tracer discordance was particularly evident at intermediate Flortaucipir TSE levels (25–50% in the MTL), where around one-quarter of participants were classified into the same MK6240 category. While spatial extent metrics offer valuable complementary information to SUVR within a single-tracer context, raw TSE values, like SUVR, should not be treated as directly interchangeable across tracers without prespecified tracer-specific calibration.

Our finding that SUVR continues to increase for MK6240 but virtually plateaus for Flortaucipir near the point of TSE saturation may have implications for detecting longitudinal tau accumulation. The larger IQR of MK6240 SUVR (MTL IQR = 1.056 vs Flortaucipir IQR = 0.240) confirms greater data dispersion among individuals with advanced tau burden, whereas the narrower IQR of Flortaucipir is consistent with a ceiling effect and SUVR signal compression near saturation. Because longitudinal studies rely on detecting within-individual SUVR change over time, a compressed dynamic range reduces statistical power and could attenuate observed rates of accumulation, particularly in early affected regions such as the entorhinal cortex and amygdala where tau pathology reaches saturation relatively early in the disease course [34, 35]. MK6240’s larger dynamic range at high tau loads suggests it may be better suited to capture the continued rate of local tau deposition in such regions, which is relevant for both studying the natural trajectory of AD and clinical trials that use changes in tau PET accumulation as an outcome measure [31].

The strengths of this study include the largest head-to-head within-individual design, in which all participants underwent both tau PET tracers within a short interval. This mitigates between-individuals variability and provides a uniquely controlled framework for direct tracer comparison. However, several limitations should be acknowledged. The design does not allow direct assessment of longitudinal trajectories of TSE or whether the inter-tracer differences observed here remain stable or change over time. Additionally, while the voxel-wise thresholds used to derive TSE were based on established methodological conventions from prior publications, the choice of threshold is inherently arbitrary, and alternative cutoffs could influence the magnitude of inter-tracer differences and the classification of individuals into TSE bins. Nevertheless, group ordering and the direction of MK6240–flortaucipir differences were preserved across alternative thresholds. Alternative reference populations or voxelwise modeling approaches could also produce different absolute TSE estimates, and our conclusions apply to the prespecified thresholding framework used here. Because TSE and SUVR are derived from the same PET images, these analyses do not establish that TSE provides incremental clinical value beyond SUVR or is generally superior across regions, tracers, or outcomes. Rather, they suggest that TSE and SUVR may provide complementary information in selected disease-stage and region-specific contexts. Finally, the absence of postmortem validation means that TSE-positive voxels cannot be confirmed as true tau pathology or directly linked to the underlying neuropathological tau deposition; autopsy-confirmed studies will be needed to establish the histopathological basis of the tracer-specific differences observed here.

Together, these findings highlight both the promise of spatial extent metrics as clinically and biologically informative measures of tau burden and the importance of tracer-specific calibration for their reliable implementation in research and clinical settings.

## Data Availability

Requests for raw or processed data and study materials may be sent to the corresponding author. Investigators and participating institutions will review each request for intellectual property or confidentiality concerns. De-identified data will be shared with qualified academic researchers seeking to reproduce the methods and findings reported in this article. Data and materials will be provided under a material transfer agreement.

## Acknowledgements

This work would not have been possible without the generosity of all the individuals who volunteered to participate in the HEAD study. We are grateful to all study participants, their families, and caregivers for their time, trust, and commitment to this research. We extend our appreciation to the clinical and research staff at the participating sites for their dedication to participant recruitment, study coordination, and data collection. We also thank all referring clinicians and community partners for their support of recruitment at all sites. We gratefully acknowledge the National Alzheimer’s Coordinating Center (NACC) for permission to utilize the Uniform Data Set in this study. We acknowledge the Laboratory of Neuroimaging (LONI) for providing the imaging & data repository, and the National Centralized Repository for Alzheimer’s Disease and Related Dementias (NCRAD) for providing the biological sample repository and APOE genotyping in this study. Finally, we extend our thanks to Lantheus/Cerveau Technologies and Avid Radiopharmaceuticals for enabling use of the MK6240 and Flortaucipir radiotracers, respectively, in this study.

## Funding sources

The HEAD study was funded by the National Institute on Aging (R01AG073267). GP is supported by the Alzheimer’s Association (24AARFD-1243899). BB is supported by the Alzheimer’s Association (AARFD-22-974627). PCLF is supported by the Alzheimer’s Association (AARFD-22-923814). GB-N is supported by the Alzheimer’s Association (AARF-D-23-1150249). PR-N receives funding from the Weston Brain Institute, Canadian Institutes of Health Research Fonds de Recherche du Québec – Santé (FRQS), Colin J. Adair Charitable Foundation. JF reports grants from the Fondo de Investigaciones Sanitario, Carlos III Health Institute (INT21/00073, and PI23/01786) and the Centro de Investigación Biomédica en Red sobre Enfermedades Neurodegenerativas (CIBERNED) Program 1, jointly funded by Fondo Europeo de Desarrollo Regional, Unión Europea, Una manera de hacer Europa. JF was also supported by the National Institutes of Health grants (1R01AG056850-01A1; R21AG056974, R01AG061566, 1R01AG081394-01 and 1R61AG066543-01), the Department de Salut de la Generalitat de Catalunya (SLT006/17/00119), Fundación Tatiana Pérez de Guzmán el Bueno (IIBSP-DOW-2020-151). He was also supported by Horizon2020–Research and Innovation Framework Programme from the European Union (H2020-SC1-BHC-2018-2020). DNS-M receives research support from the NIH (K23-AG076960). Development of the NACC Uniform Data Set was funded by the National Institutes of Health through the National Institute on Aging (Cooperative Agreement U01AG016976). The National Centralized Repository for Alzheimer’s Disease and Related Dementias (NCRAD) is supported by the National Institutes of Health through the National Institute on Aging (U24AG021886). We used artificial intelligence-assisted tools to improve grammar and readability, without altering the scientific content or meaning.

## Conflicts of interest

TAP serves on a Data Safety Monitoring Board for National Institutes of Health (NIH) award 5 R01 AG084834 based at the University of Cincinnati. He also has served on a scientific advisory board at Johnson & Johnson. TAP, BB, GP, PCFL, and GB-N have received support to attend meetings from the Alzheimer’s Association. PR-N serves as Advisory Board member from Novonordisk, Eisai and Ely Lilly. JF reported receiving personal fees for service on the advisory boards, adjudication committees or speaker honoraria from AC Immune, Adamed, Alzheon, Biogen, Eisai, Esteve, Fujirebio, Ionis, Laboratorios Carnot, Life Molecular Imaging, Lilly, Novo Nordisk, Perha, Roche, Zambón and outside the submitted work. He reports holding a patent for markers of synaptopathy in neurodegenerative disease (licensed to ADx, EPI8382175.0). DNS-M received a travel award from the Society of Nuclear Medicine and Molecular Imaging and an honorarium for grant review from the Alzheimer’s Drug Discovery Foundation. He serves on a research council for the Alzheimer’s Association and on a Data Monitoring Committee for Eli Lilly and Company. HO serves on a Data Safety Monitoring Board for National Institutes of Health (NIH) award 5 R01 AG068062 based at Massachusetts General Hospital. DLT participated on a Data Safety Monitoring board for DSMS REMBRANDT study. TKK has consulted for Quanterix Corporation, SpearBio Inc., Neurogen Biomarking LLC., and Alzheon, has served on advisory boards for Siemens Healthineers and Neurogen Biomarking LLC., outside the submitted work. Over the last three years, he has received in-kind research support from Janssen Research Laboratories, SpearBio Inc., and Alamar Biosciences, as well as meeting travel support from the Alzheimer’s Association and Neurogen Biomarking LLC., outside the submitted work. TKK has received royalties from Bioventix for the transfer of specific antibodies and assays to third party organizations. TKK is inventor on several patents and provisional patents regarding biofluid biomarker methods, targets and reagents/compositions, that may generate income for the institution and/or self should they be licensed and/or transferred to another organization. All other authors declare no competing interests.

## Consent Statement

All participants provided written informed consent.

## Notes

### Clinical Trial

NCT05361382

### Author Declarations

The Advarra Institutional Review Board served as the central IRB of record for the participating sites and gave ethical approval for this work (Protocol ID: Pro00061365).

