## Supplemental Material for "Detection of tau spatial extent using MK6240 and flortaucipir PET"

### **Supplemental Methods**

#### **Scanner harmonization and phantom calibration**

To minimize the influence of differing intrinsic spatial resolution across PET scanners on regional SUVR estimates, the point-spread function (PSF) of each scanner was characterized using site-specific phantom and used to harmonize all PET images to a common target resolution of 8 mm FWHM. For each scanner, the additional Gaussian smoothing kernel required to reach this target was computed by quadrature subtraction of the intrinsic PSF from the target FWHM, and converted to the corresponding Gaussian sigma using the standard analytical relationship between sigma and FWHM.

### **Supplemental Figures**

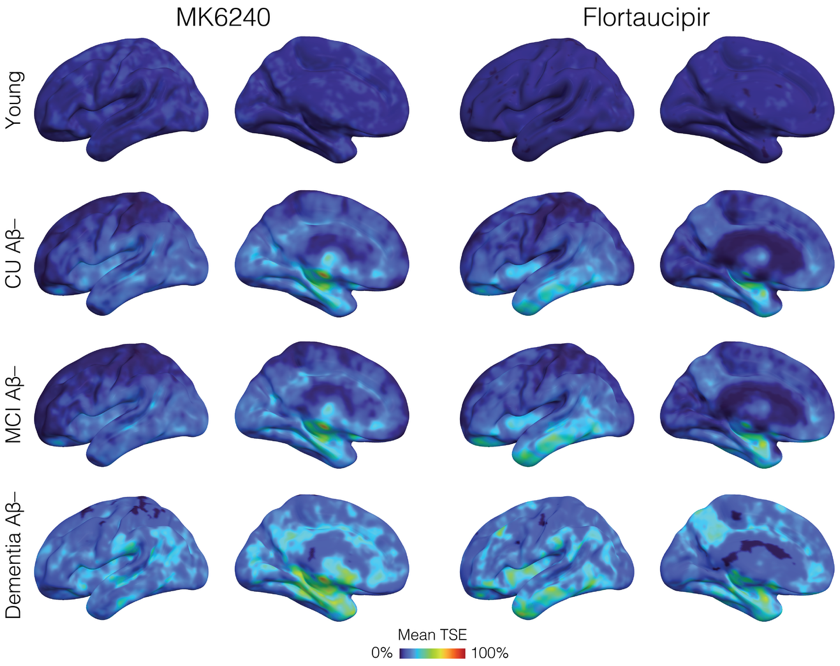

**Supplemental Figure 1. Topographical distribution of abnormal tau spatial extent for MK6240 and Flortaucipir in amyloid-β negative (Aβ-) individuals.** CU = cognitively unimpaired older adults; MCI = mild cognitive impairment; Young = individuals between 18-28 years old.

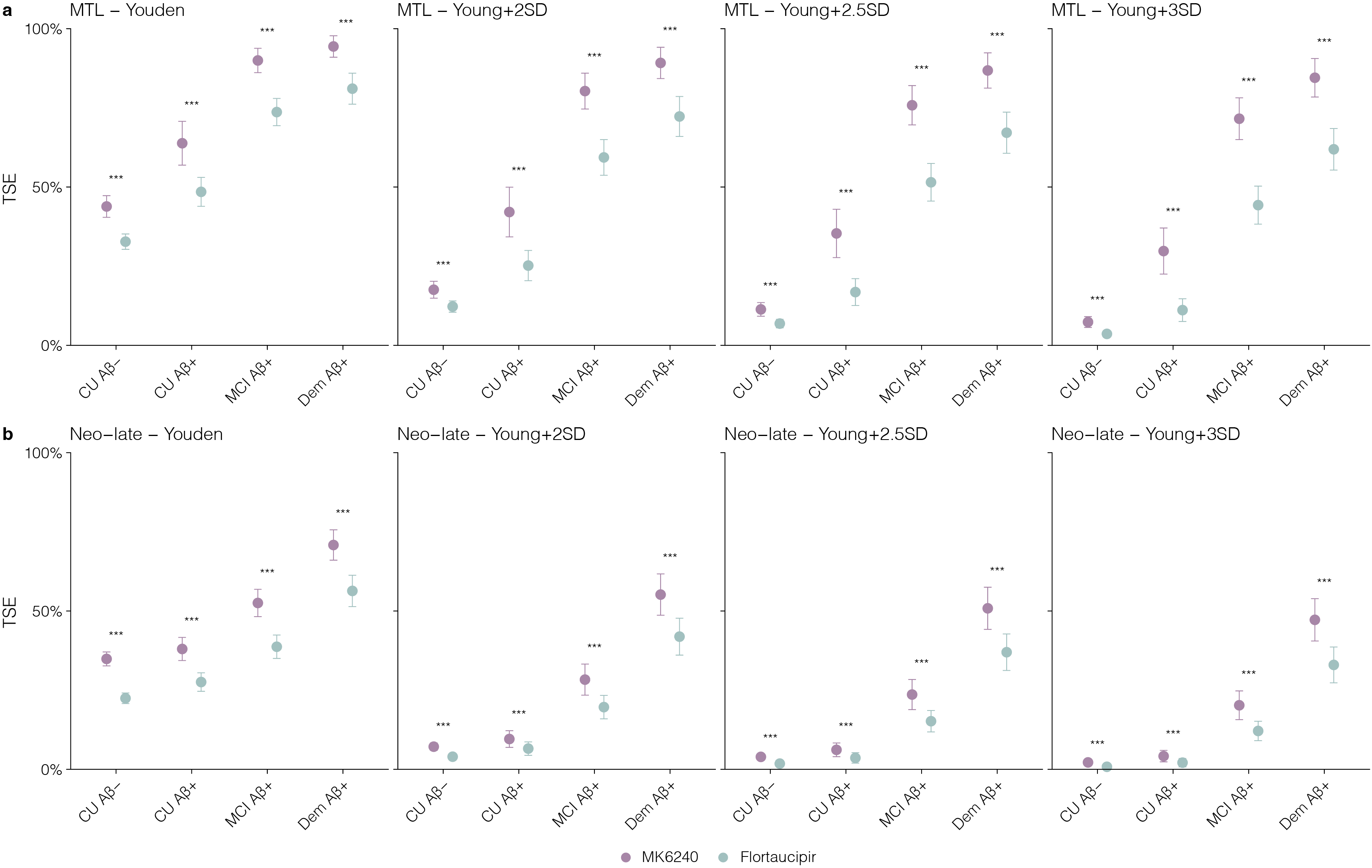

**Supplemental Figure 2. Tau spatial extent (TSE) by clinical group is consistent across voxel-classification thresholds.** Mean TSE with 95% confidence intervals for MK6240 (purple) and flortaucipir (teal) across four clinical groups (CU Aβ−, CU Aβ+, MCI Aβ+, Dementia Aβ+). The medial temporal lobe (MTL) is shown in the top row and the neocortical late composite (Neo-late) in the bottom row. Each column applies a different voxel-level positivity threshold used to classify individual voxels as abnormal before computing TSE, from left to right a Youden-optimized threshold, and young-reference mean plus 2, 2.5, and 3 standard deviations. Asterisks denote a significant difference between tracers within each clinical group by paired Wilcoxon signed-rank test (***p < 0.001).

### **Supplemental Tables**

**Supplemental Table 1. Scanner models, reconstruction, and correction parameters**

| Site | Scanner model | Recon. algorithm | Iter. × subsets | Voxel size (mm) | Correction |
| --- | --- | --- | --- | --- | --- |
| Mayo | GE Discovery 690 | VPHD/VPFX | 4 × 24 | 1.33×1.33×1.96 | CT-based |
| Mayo | GE Discovery MI | VPHD/VPFX | 6 × 16 | 1.33×1.33×2.79 | CT-based |
| Mayo | Siemens Biograph Vision 600 | TOF-OSEM | 8 × 5 | 0.825×0.825×1.65 | CT-based |
| McGill | GE Discovery 690 | VPHD/VPFX | 4 × 24 | 1.33×1.33×1.96 | CT-based |
| McGill | Siemens HRRT | OP-OSEM-3D | 6 × 16 | 1.219×1.219×1.219 | Transmission-based |
| UCSF | Siemens Biograph Vision 600 | TOF-OSEM | 8 × 5 | 0.825×0.825×1.65 | CT-based |
| Houston | Siemens Biograph Vision 600 Edge | TOF-OSEM | 8 × 5 | 0.825×0.825×1.65 | CT-based |
| Houston | Siemens Biograph Vision Quadra Edge | TOF-OSEM | 8 × 5 | 0.825×0.825×1.65 | CT-based |
| WashU | Siemens Biograph Vision 600 Edge | TOF-OSEM | 8 × 5 | 0.825×0.825×1.65 | CT-based |
| WashU | Siemens Biograph mCT | OSEM | 4 × 24 | 1.018×1.018×2.027 | CT-based |
| Pittsburgh | Siemens Biograph mCT | OSEM | 4 × 24 | 1.018×1.018×2.027 | CT-based |
| Brown | Siemens Biograph mCT | OSEM | 4 × 24 | 1.018×1.018×2.027 | CT-based |
| LBL | Siemens Biograph mCT | OSEM | 4 × 24 | 1.018×1.018×2.027 | CT-based |

No post-reconstruction filters were applied in any site/scanner. Recon = reconstruction; Iter = iterations; CT = computed tomography; FWHM = full width at half-maximum; OSEM = ordered-subsets expectation maximization; PSF = point-spread function; SUVR = standardized uptake value ratio; TOF = time of flight; VPHD/VPFX = GE HealthCare iterative reconstruction algorithms.

#### **Supplemental Table 2. Participant characteristics.**

|  | Young (N=38) | CU elderly (N=362) | MCI (N=183) | Dementia (N=99) |
| --- | --- | --- | --- | --- |
| Age, mean (SD) | 23 (2.0) | 67 (8.3) | 71 (7.4) | 72 (8.9) |
| Sex, n (% of female) | 23 (61%) | 220 (61%) | 82 (45%) | 48 (48%) |
| APOEε4, n (% of carrier) | 11 (29 %) | 109 (30 %) | 94 (51 %) | 59 (60 %) |
| Education, years (SD) | 16 (2.0) | 16 (2.5) | 16 (2.9) | 15 (3.3) |
| MMSE, mean (SD) | 29 (1.0) | 29 (1.5) | 28 (2.0) | 21 (4.9) |
| CDR sum of boxes, mean (SD) | 0 (0) | 0.02 (0.10) | 1.7 (1.2) | 4.7 (2.5) |
| MoCA, mean (SD) | 28 (2.0) | 27 (2.2) | 25 (5.7) | 17 (9.1) |
| Centiloid, mean (SD) | -1.2 (4.2) | 15 (33) | 55 (57) | 95 (50) |
| Aβ status, n (% of positive) | 0 (0 %) | 77 (21 %) | 108 (59 %) | 87 (88 %) |

Missing: 5 APOEε4 status, 6 education years, 5 MMSE score, 1 CDR-SB. CU = cognitively unimpaired; MCI = mild cognitive impairment; SD = standard deviation; MTL = medial temporal lobe; Neo-late = late neocortical region comprising Braak V-VI; TSE = tau spatial extent; SUVR; standardized uptake value ratio.

#### **Supplemental Table 3. Mean percentage of abnormal voxels (TSE) across Braak regions by clinical group and tracer.**

| Group | Braak | Flortaucipir Mean (SD) | MK6240 Mean (SD) | P-value |
| --- | --- | --- | --- | --- |
| CU elderly Aβ– | I | 13% (22%) | 21% (30%) | < 0.001 |
|  | II | 12% (15%) | 17% (22%) | < 0.001 |
|  | III | 6% (9%) | 11% (15%) | < 0.001 |
|  | IV | 9% (11%) | 9% (12%) | 0.80714 |
|  | V | 4% (7%) | 7% (10%) | < 0.001 |
|  | VI | 2% (5%) | 7% (11%) | < 0.001 |
| CU elderly Aβ+ | I | 31% (31%) | 49% (42%) | < 0.001 |
|  | II | 25% (22%) | 41% (38%) | < 0.001 |
|  | III | 10% (12%) | 20% (22%) | < 0.001 |
|  | IV | 15% (16%) | 15% (19%) | 0.74290 |
|  | V | 7% (11%) | 10% (13%) | 0.00423 |
|  | VI | 3% (7%) | 9% (12%) | < 0.001 |
| MCI Aβ– | I | 18% (22%) | 29% (34%) | 0.00176 |
|  | II | 18% (16%) | 22% (26%) | 0.14440 |
|  | III | 8% (11%) | 14% (18%) | 0.00304 |
|  | IV | 14% (14%) | 12% (14%) | 0.21561 |
|  | V | 6% (9%) | 8% (9%) | 0.12633 |
|  | VI | 3% (6%) | 7% (9%) | < 0.001 |
| MCI Aβ+ | I | 65% (34%) | 82% (32%) | < 0.001 |
|  | II | 59% (30%) | 80% (30%) | < 0.001 |
|  | III | 35% (27%) | 52% (33%) | < 0.001 |
|  | IV | 38% (27%) | 46% (35%) | < 0.001 |
|  | V | 22% (21%) | 30% (27%) | < 0.001 |
|  | VI | 12% (16%) | 22% (23%) | < 0.001 |
| Dementia Aβ– | I | 21% (30%) | 34% (39%) | 0.18514 |
|  | II | 25% (25%) | 30% (34%) | 0.68908 |
|  | III | 10% (11%) | 16% (17%) | 0.30656 |
|  | IV | 15% (15%) | 14% (15%) | 0.72408 |
|  | V | 10% (10%) | 11% (12%) | 1.00000 |
|  | VI | 8% (10%) | 12% (14%) | 0.16817 |
| Dementia Aβ+ | I | 80% (30%) | 92% (22%) | < 0.001 |
|  | II | 71% (30%) | 89% (24%) | < 0.001 |
|  | III | 60% (30%) | 76% (28%) | < 0.001 |
|  | IV | 64% (28%) | 75% (31%) | < 0.001 |
|  | V | 46% (29%) | 58% (32%) | < 0.001 |
|  | VI | 26% (24%) | 44% (30%) | < 0.001 |

P-values from paired Wilcoxon signed-rank tests comparing MK6240 vs Flortaucipir. CU: cognitively unimpaired; MCI: mild cognitive impairment; TSE = tau spatial extent; SD = standard deviation.

#### **Supplemental Table 4. Comparison of Flortaucipir and MK6240 spatial extent (TSE) by clinical group.**

| Region | Group | Flortaucipir Mean (SD) | MK6240 Mean (SD) | P-value |
| --- | --- | --- | --- | --- |
| MTL | CU elderly Aβ– | 0.12 (0.15) | 0.18 (0.23) | < 0.001 |
|  | CU elderly Aβ+ | 0.25 (0.23) | 0.42 (0.38) | < 0.001 |
|  | MCI Aβ+ | 0.59 (0.30) | 0.80 (0.30) | < 0.001 |
|  | Dementia Aβ+ | 0.72 (0.30) | 0.89 (0.24) | < 0.001 |
| Neo-late | CU elderly Aβ– | 0.04 (0.06) | 0.07 (0.10) | < 0.001 |
|  | CU elderly Aβ+ | 0.07 (0.10) | 0.10 (0.13) | < 0.001 |
|  | MCI Aβ+ | 0.20 (0.20) | 0.28 (0.26) | < 0.001 |
|  | Dementia Aβ+ | 0.42 (0.28) | 0.55 (0.31) | < 0.001 |

P-values from paired Wilcoxon signed-rank tests. CU: cognitively unimpaired; MCI: mild cognitive impairment; SD = standard deviation; MTL = medial temporal lobe.

#### **Supplemental Table 5. Comparison of Flortaucipir and MK6240 spatial extent (TSE) across brain regions stratified by Centiloid bins.**

| Region | Centiloid bin | Flortaucipir Mean (SD) | MK6240 Mean (SD) | P-value |
| --- | --- | --- | --- | --- |
| MTL | < 10 | 0.12 (0.15) | 0.17 (0.23) | < 0.001 |
|  | 10–30 | 0.23 (0.20) | 0.32 (0.32) | 0.02820 |
|  | 30–50 | 0.28 (0.24) | 0.47 (0.37) | < 0.001 |
|  | 50–75 | 0.43 (0.31) | 0.68 (0.36) | < 0.001 |
|  | 75–100 | 0.58 (0.31) | 0.78 (0.33) | < 0.001 |
|  | > 100 | 0.72 (0.28) | 0.91 (0.21) | < 0.001 |
| Neo-late | < 10 | 0.04 (0.06) | 0.07 (0.10) | < 0.001 |
|  | 10–30 | 0.09 (0.13) | 0.11 (0.14) | 0.04168 |
|  | 30–50 | 0.07 (0.12) | 0.09 (0.11) | 0.05589 |
|  | 50–75 | 0.15 (0.24) | 0.22 (0.29) | < 0.001 |
|  | 75–100 | 0.19 (0.21) | 0.26 (0.26) | < 0.001 |
|  | > 100 | 0.36 (0.26) | 0.49 (0.30) | < 0.001 |

P-values from paired Wilcoxon signed-rank tests. SD = standard deviation; MTL = medial temporal lobe.

#### **Supplemental Table 6. Comparison of Flortaucipir and MK6240 spatial extent (TSE) across brain regions stratified by CDR-SB bins.**

| Region | CDR-SB bin | Flortaucipir Mean (SD) | MK6240 Mean (SD) | P-value |
| --- | --- | --- | --- | --- |
| MTL | 0 | 0.14 (0.18) | 0.21 (0.28) | < 0.001 |
|  | 0.5–1 | 0.35 (0.29) | 0.47 (0.38) | < 0.001 |
|  | 1.5–2.5 | 0.47 (0.35) | 0.63 (0.40) | < 0.001 |
|  | 3–4 | 0.64 (0.32) | 0.79 (0.32) | < 0.001 |
|  | >= 4.5 | 0.67 (0.32) | 0.82 (0.32) | < 0.001 |
| Neo-late | 0 | 0.04 (0.07) | 0.07 (0.10) | < 0.001 |
|  | 0.5–1 | 0.11 (0.15) | 0.15 (0.19) | < 0.001 |
|  | 1.5–2.5 | 0.18 (0.22) | 0.24 (0.27) | < 0.001 |
|  | 3–4 | 0.27 (0.23) | 0.38 (0.30) | < 0.001 |
|  | >= 4.5 | 0.41 (0.28) | 0.54 (0.33) | < 0.001 |

P-values from paired Wilcoxon signed-rank tests. SD = standard deviation; MTL = medial temporal lobe.

#### **Supplemental Table 7. Percentage of participants exceeding TSE positivity thresholds by tracer and region.**

| Region | Threshold | Flortaucipir N (%) | MK6240 N (%) | McNemar P |
| --- | --- | --- | --- | --- |
| MTL | ≥ 25% | 280 (41.1%) | 337 (49.4%) | < 0.001 |
|  | ≥ 50% | 162 (23.8%) | 248 (36.4%) | < 0.001 |
|  | ≥ 75% | 105 (15.4%) | 197 (28.9%) | < 0.001 |
| Neo-late | ≥ 25% | 110 (16.1%) | 150 (22.0%) | < 0.001 |
|  | ≥ 50% | 48 (7.0%) | 72 (10.6%) | < 0.001 |
|  | ≥ 75% | 14 (2.1%) | 38 (5.6%) | < 0.001 |

P-values from McNemar tests comparing paired positivity rates. TSE = tau spatial extent; MTL = medial temporal lobe.

#### **Supplemental Table 8. Sex-stratified paired comparisons of flortaucipir and MK6240 TSE.**

| Region | Male, median ΔTSE  (P-value) | Female, median ΔTSE  (P-value) | Difference by sex P-value |
| --- | --- | --- | --- |
| MTL | 0.080 (P <0.001) | 0.039 (P <0.001) | 0.400 |
| Neo-late | 0.017 (P <0.001) | 0.026 (P <0.001) | 0.973 |

Within-sex P values are from paired Wilcoxon tests. Between-sex P values are from Welch two-sample t-tests comparing participant-level ΔTSE. ΔTSE = MK6240 − flortaucipir. TSE = tau spatial extent; MTL = medial temporal lobe.

#### **Supplemental Table 9. Prevalence ratios (MK6240 vs Flortaucipir) from Poisson regression.**

| TSE Threshold | Region | Group | PR (95% CI) | P-value |
| --- | --- | --- | --- | --- |
| ≥ 25% | MTL | Cognitively unimpaired | 1.42 (1.04–1.93) | 0.02558 |
|  | MTL | Cognitively impaired | 1.11 (1.04–1.19) | 0.00175 |
|  | Neo-late | Cognitively unimpaired | 2.25 (0.74–6.84) | 0.15293 |
|  | Neo-late | Cognitively impaired | 1.23 (1.04–1.46) | 0.01501 |
| ≥ 50% | MTL | Cognitively unimpaired | 2.36 (1.40–3.98) | 0.00134 |
|  | MTL | Cognitively impaired | 1.22 (1.11–1.34) | < 0.001 |
|  | Neo-late | Cognitively unimpaired | 0.50 (0.06–4.27) | 0.52630 |
|  | Neo-late | Cognitively impaired | 1.52 (1.12–2.07) | 0.00706 |

TSE = tau spatial extent; CI = confidence interval; PR = prevalence ratio; MTL = medial temporal lobe.

#### **Supplemental Table 10. Logistic regression parameters for P(TSE ≥ threshold) ~ Centiloid.**

| TSE Threshold | Region | Tracer | % Tau+ | OR per CL unit (95% CI) | CL50 |
| --- | --- | --- | --- | --- | --- |
| ≥ 25% | MTL | MK6240 | 52.1% | 1.0370 (1.0303–1.0437) | 25.7 |
|  | MTL | Flortaucipir | 43.5% | 1.0285 (1.0238–1.0333) | 45.2 |
|  | Neo-late | MK6240 | 23.2% | 1.0302 (1.0255–1.0350) | 95.3 |
|  | Neo-late | Flortaucipir | 17.1% | 1.0288 (1.0238–1.0339) | 117.6 |
| ≥ 50% | MTL | MK6240 | 38.7% | 1.0368 (1.0313–1.0424) | 51.0 |
|  | MTL | Flortaucipir | 25.2% | 1.0368 (1.0313–1.0423) | 84.4 |
|  | Neo-late | MK6240 | 11.1% | 1.0340 (1.0271–1.0410) | 136.4 |
|  | Neo-late | Flortaucipir | 7.4% | 1.0261 (1.0197–1.0326) | 170.6 |

OR = odds ratio per one-unit Centiloid increase; CL50 = Centiloid at P(tau+) = 50%. TSE = tau spatial extent; CL = Centiloid; CI = confidence interval; MTL = medial temporal lobe.

#### **Supplemental Table 11. Additional tau-positive cases detected with MK6240 vs Flortaucipir.**

| TSE Threshold | Region | Group | ARR | NND (95% CI) |
| --- | --- | --- | --- | --- |
| ≥ 25% | MTL | CU Aβ+ | 0.1459 | 6.9 (4.2–17.6) |
|  |  | MCI Aβ+ | 0.1022 | 9.8 (5.7–21.6) |
|  |  | Dementia Aβ+ | 0.0821 | 12.2 (7.2–28.7) |
|  | Neo-late | MCI Aβ+ | 0.1107 | 9.0 (5.4–27.0) |
|  |  | Dementia Aβ+ | 0.1132 | 8.8 (5.1–21.5) |
| ≥ 50% | MTL | CU Aβ+ | 0.2393 | 4.2 (3.0–6.8) |
|  |  | MCI Aβ+ | 0.2121 | 4.7 (3.4–7.2) |
|  |  | Dementia Aβ+ | 0.0922 | 10.9 (6.1–28.7) |
|  | Neo-late | MCI Aβ+ | 0.1104 | 9.1 (6.0–18.0) |
|  |  | Dementia Aβ+ | 0.1403 | 7.1 (4.3–17.2) |

ARR = absolute risk reduction; NND = number needed to detect. 95% CI from 1000 bootstrap resamples. CU = cognitively unimpaired; MCI = mild cognitive impairment; TSE = tau spatial extent; CI = confidence interval; MTL = medial temporal lobe.

#### **Supplemental Table 12. Tracer dynamic range in regions of spatial saturation.**

| Region | Tracer | N (Saturated) | IQR | SD | Min SUVR | Max SUVR |
| --- | --- | --- | --- | --- | --- | --- |
| MTL | Flortaucipir | 105 | 0.240 | 0.184 | 1.42 | 2.33 |
|  | MK6240 | 197 | 1.056 | 0.684 | 1.08 | 4.51 |
| Neo-late | Flortaucipir | 14 | 0.183 | 0.359 | 1.66 | 3.00 |
|  | MK6240 | 38 | 0.918 | 0.873 | 1.66 | 5.08 |

Saturation defined as ≥ 75% of region affected (TSE). IQR = interquartile range of SUVR among saturated participants. SD = standard deviation; TSE = tau spatial extent; SUVR = standardized uptake value ratio; MTL = medial temporal lobe.

#### **Supplemental Table 13. Spearman correlations of TSE and SUVR with clinical and biomarker outcomes, with Steiger z-test.**

| Region | Tracer | ρ (TSE) | ρ (SUVR) | Steiger Z | P-value |
| --- | --- | --- | --- | --- | --- |
| **Memory composite score** |  |  |  |  |  |
| MTL | MK6240 | -0.507 | -0.502 | 0.65 | 0.51275 |
| MTL | Flortaucipir | -0.489 | -0.458 | 2.69 | 0.00704 |
| Neo-late | MK6240 | -0.435 | -0.330 | 6.71 | < 0.001 |
| Neo-late | Flortaucipir | -0.438 | -0.256 | 8.23 | < 0.001 |
| **Centiloid** |  |  |  |  |  |
| MTL | MK6240 | 0.648 | 0.639 | 1.31 | 0.19155 |
| MTL | Flortaucipir | 0.631 | 0.591 | 3.90 | < 0.001 |
| Neo-late | MK6240 | 0.509 | 0.429 | 5.47 | < 0.001 |
| Neo-late | Flortaucipir | 0.547 | 0.411 | 6.69 | < 0.001 |
| **pTau217** |  |  |  |  |  |
| MTL | MK6240 | 0.632 | 0.628 | 0.47 | 0.63667 |
| MTL | Flortaucipir | 0.596 | 0.572 | 2.23 | 0.02547 |
| Neo-late | MK6240 | 0.425 | 0.332 | 5.94 | < 0.001 |
| Neo-late | Flortaucipir | 0.422 | 0.249 | 7.72 | < 0.001 |
| **CDR-SB** |  |  |  |  |  |
| MTL | MK6240 | 0.563 | 0.551 | 1.57 | 0.11601 |
| MTL | Flortaucipir | 0.568 | 0.525 | 3.94 | < 0.001 |
| Neo-late | MK6240 | 0.483 | 0.391 | 6.12 | < 0.001 |
| Neo-late | Flortaucipir | 0.502 | 0.341 | 7.57 | < 0.001 |
| **MoCA** |  |  |  |  |  |
| MTL | MK6240 | -0.469 | -0.460 | 1.16 | 0.24543 |
| MTL | Flortaucipir | -0.450 | -0.404 | 3.90 | < 0.001 |
| Neo-late | MK6240 | -0.398 | -0.320 | 4.92 | < 0.001 |
| Neo-late | Flortaucipir | -0.410 | -0.253 | 7.03 | < 0.001 |

Steiger z-test on ranked data. Positive z indicates |ρ(TSE)| > |ρ(SUVR)|. TSE = tau spatial extent; SUVR = standardized uptake value ratio; MTL = medial temporal lobe; CDR-SB = clinical dementia rating - sum of boxes; MoCA = montreal cognitive assessment;

#### **Supplemental Table 14. Cohen's d for clinical group separations: TSE vs SUVR with Steiger z-test.**

| Comparison | Tracer | d TSE (95% CI) | d SUVR (95% CI) | Steiger Z | P-value |
| --- | --- | --- | --- | --- | --- |
| **MTL** | | | | | |
| CU Aβ+ vs CU Aβ− | MK6240 | 0.91 (0.61–1.26) | 0.95 (0.69–1.21) | -0.59 | 0.55600 |
| CU Aβ+ vs CU Aβ− | Flortaucipir | 0.75 (0.50–1.04) | 0.71 (0.46–0.99) | 0.74 | 0.46205 |
| MCI Aβ+ vs CU Aβ− | MK6240 | 2.52 (2.12–3.03) | 2.17 (1.90–2.51) | 2.52 | 0.01186 |
| MCI Aβ+ vs CU Aβ− | Flortaucipir | 2.31 (1.96–2.76) | 2.05 (1.77–2.38) | 3.59 | < 0.001 |
| Dem Aβ+ vs CU Aβ− | MK6240 | 3.14 (2.65–3.78) | 3.37 (2.88–4.04) | -1.25 | 0.21281 |
| Dem Aβ+ vs CU Aβ− | Flortaucipir | 3.05 (2.53–3.73) | 2.83 (2.38–3.40) | 2.06 | 0.03915 |
| MCI Aβ+ vs CI Aβ− | MK6240 | 1.95 (1.55–2.60) | 1.51 (1.32–1.81) | 2.81 | 0.00495 |
| MCI Aβ+ vs CI Aβ− | Flortaucipir | 1.61 (1.30–2.03) | 1.45 (1.22–1.75) | 2.13 | 0.03312 |
| Dem Aβ+ vs CI Aβ− | MK6240 | 2.56 (2.03–3.28) | 2.32 (1.96–2.80) | 1.24 | 0.21653 |
| Dem Aβ+ vs CI Aβ− | Flortaucipir | 2.17 (1.72–2.82) | 1.97 (1.64–2.43) | 1.95 | 0.05131 |
| **Neo-late** | | | | | |
| CU Aβ+ vs CU Aβ− | MK6240 | 0.23 (-0.02–0.49) | 0.20 (-0.04–0.43) | 0.47 | 0.64086 |
| CU Aβ+ vs CU Aβ− | Flortaucipir | 0.34 (0.09–0.59) | 0.29 (0.07–0.52) | 0.61 | 0.54111 |
| MCI Aβ+ vs CU Aβ− | MK6240 | 1.31 (1.05–1.60) | 0.88 (0.70–1.06) | 6.27 | < 0.001 |
| MCI Aβ+ vs CU Aβ− | Flortaucipir | 1.33 (1.10–1.60) | 0.79 (0.58–1.02) | 7.34 | < 0.001 |
| Dem Aβ+ vs CU Aβ− | MK6240 | 2.75 (2.29–3.31) | 1.61 (1.38–1.91) | 9.24 | < 0.001 |
| Dem Aβ+ vs CU Aβ− | Flortaucipir | 2.58 (2.19–3.05) | 1.55 (1.30–1.89) | 10.39 | < 0.001 |
| MCI Aβ+ vs CI Aβ− | MK6240 | 1.00 (0.79–1.24) | 0.69 (0.55–0.85) | 3.55 | < 0.001 |
| MCI Aβ+ vs CI Aβ− | Flortaucipir | 0.85 (0.63–1.08) | 0.63 (0.42–0.85) | 2.85 | 0.00439 |
| Dem Aβ+ vs CI Aβ− | MK6240 | 2.05 (1.69–2.52) | 1.14 (0.97–1.35) | 6.45 | < 0.001 |
| Dem Aβ+ vs CI Aβ− | Flortaucipir | 1.74 (1.44–2.08) | 1.11 (0.91–1.38) | 5.92 | < 0.001 |

Steiger z via d-to-r conversion. Bootstrap 95% CI from 1000 resamples. CU = cognitively unimpaired; MCI = mild cognitive impairment; Dem = dementia; TSE = tau spatial extent; SUVR = standardized uptake value ratio; MTL = medial temporal lobe.

#### **Supplemental Table 15. SUVR positivity rates within TSE bins by tracer and region.**

| Region | Tracer | TSE Bin | SUVR+ Rate |
| --- | --- | --- | --- |
| MTL | MK6240 | 0–25% | 0.0% |
|  |  | 25–50% | 16.9% |
|  |  | > 50% | 99.6% |
|  | Flortaucipir | 0–25% | 0.0% |
|  |  | 25–50% | 10.2% |
|  |  | > 50% | 100.0% |
| Neo-late composite | MK6240 | 0–25% | 0.0% |
|  |  | 25–50% | 17.9% |
|  |  | > 50% | 100.0% |
|  | Flortaucipir | 0–25% | 0.0% |
|  |  | 25–50% | 0.0% |
|  |  | > 50% | 100.0% |

TSE = tau spatial extent; SUVR = standardized uptake value ratio; MTL = medial temporal lobe.

#### **Supplemental Table 16. Centiloid and plasma pTau-217 in participants within 0–25% vs 25–50% TSE bins.**

| Region | Tracer | Outcome | 0-25% TSE: Mean (SD) | 25-50% TSE: Mean (SD) | Wilcoxon P |
| --- | --- | --- | --- | --- | --- |
| MTL | MK6240 | Centiloid | 6.53 (20.81) | 22.67 (39.14) | < 0.001 |
|  |  | pTau-217 | 0.34 (0.21) | 0.45 (0.32) | 0.00791 |
|  | Flortaucipir | Centiloid | 11.36 (27.71) | 31.37 (45.60) | < 0.001 |
|  |  | pTau-217 | 0.38 (0.26) | 0.53 (0.34) | < 0.001 |
| Neo-late | MK6240 | Centiloid | 19.02 (36.08) | 76.18 (58.89) | < 0.001 |
|  |  | pTau-217 | 0.45 (0.32) | 0.87 (0.56) | < 0.001 |
|  | Flortaucipir | Centiloid | 23.10 (41.13) | 96.96 (55.91) | < 0.001 |
|  |  | pTau-217 | 0.48 (0.35) | 1.05 (0.64) | < 0.001 |

P-values from Wilcoxon rank-sum tests. TSE = tau spatial extent; SD = standard deviation; MTL = medial temporal lobe.

#### **Supplemental Table 17. Odds of Aβ and plasma pTau-217 positivity in the 25–50% TSE bin relative to 0–25%, restricted to SUVR-negative individuals.**

| Tracer | Outcome | OR (95% CI) | P-value |
| --- | --- | --- | --- |
| MK6240 | Aβ PET+ | 3.55 (1.94–6.51) | <0.001 |
| MK6240 | pTau-217+ | 3.14 (1.77–5.57) | <0.001 |
| Flortaucipir | Aβ PET+ | 3.04 (1.89–4.88) | <0.001 |
| Flortaucipir | pTau-217+ | 2.92 (1.84–4.65) | <0.001 |

P-values from Wald z-tests. Wald 95% CIs from logistic regression. TSE = tau spatial extent; SUVR = standardized uptake value ratio; OR = odds ratio; CI = confidence interval. Aβ = amyloid-β;TSE = tau spatial extent; SUVR = standardized uptake value ratio.

#### **Supplemental Table 18. Cross-tracer TSE agreement statistics.**

| Region | Spearman ρ | ρ P-value | Mean Diff  (MK6240 - Flortaucipir) | SD of Diff | MAE | Wilcoxon P (offset) |
| --- | --- | --- | --- | --- | --- | --- |
| MTL | 0.877 | < 0.001 | 0.106 | 0.190 | 0.147 | < 0.001 |
| Neo-late | 0.789 | < 0.001 | 0.050 | 0.105 | 0.074 | < 0.001 |

Paired Wilcoxon test for systematic offset. MAE = mean absolute error. Positive mean diff = MK6240 > Flortaucipir. TSE = tau spatial extent; MTL = medial temporal lobe.

#### **Supplemental Table 19. Cross-classification of TSE between Flortaucipir and MK6240.**

| Region | Flortaucipir Bin | MK6240 Lower (%) | MK6240 Same (%) | MK6240 Higher (%) |
| --- | --- | --- | --- | --- |
| MTL | 0–25% | 0.0% | 79.4% | 20.6% |
|  | 25–50% | 21.2% | 25.4% | 53.4% |
|  | 50–75% | 7.0% | 10.5% | 82.5% |
|  | 75–100% | 0.0% | 100.0% | 0.0% |
| Neo-late | 0–25% | 0.0% | 90.9% | 9.1% |
|  | 25–50% | 16.1% | 48.4% | 35.5% |
|  | 50–75% | 8.8% | 23.5% | 67.6% |
|  | 75–100% | 0.0% | 100.0% | 0.0% |

TSE = tau spatial extent; MTL = medial temporal lobe.

#### **Supplemental Table 20. Cross-classification of TSE between MK6240 and Flortaucipir.**

| Region | MK6240 Bin | Flortaucipir Lower (%) | Flortaucipir Same (%) | Flortaucipir Higher (%) |
| --- | --- | --- | --- | --- |
| MTL | 0–25% | 0.0% | 92.5% | 7.5% |
|  | 25–50% | 62.9% | 33.7% | 3.4% |
|  | 50–75% | 88.2% | 11.8% | 0.0% |
|  | 75–100% | 46.7% | 53.3% | 0.0% |
| Neo-late | 0–25% | 0.0% | 97.7% | 2.3% |
|  | 25–50% | 60.3% | 38.5% | 1.3% |
|  | 50–75% | 76.5% | 23.5% | 0.0% |
|  | 75–100% | 63.2% | 36.8% | 0.0% |

TSE = tau spatial extent; MTL = medial temporal lobe.

#### **Supplemental Table 21. Cross-tracer bias and proportional bias**

| Region | Metric | Mean bias (95% CI) | 95% LoA | Proportional-bias slope  (P-value) |
| --- | --- | --- | --- | --- |
| MTL | TSE | 10.6 (9.1–12.0) | −26.7 to 47.9 | 0.235 (<0.001) |
| MTL | SUVR | 0.003 (−0.029–0.036) | −0.850 to 0.856 | 0.892 (<0.001) |
| Neo-late | TSE | 5.0 (4.3–5.9) | −15.6 to 25.7 | 0.245 (<0.001) |
| Neo-late | SUVR | −0.062 (−0.083–−0.037) | −0.653 to 0.529 | 0.887 (<0.001) |

Bias is MK6240 – flortaucipir. TSE bias is expressed in percentage; SUVR bias is expressed in SUVR units. Limits of agreement are bias ±1.96 SD of paired differences. Proportional bias was assessed by regressing paired differences on paired means. TSE = tau spatial extent; SUVR = standardized uptake value ratio; MTL = medial temporal lobe; CI = confidence interval.

#### **Supplemental Table 22. Cross-tracer concordance and calibration**

| Region | Metric | CCC (95% CI) | Deming intercept (95% CI) | Deming slope (95% CI) |
| --- | --- | --- | --- | --- |
| MTL | TSE | 0.824 (0.796–0.850) | 0.022 (0.008–0.037) | 1.285 (1.241–1.336) |
| MTL | SUVR | 0.628 (0.603–0.653) | −2.045 (−2.224–−1.874) | 2.686 (2.536–2.839) |
| Neo-late | TSE | 0.858 (0.825–0.884) | 0.016 (0.009–0.022) | 1.295 (1.244–1.361) |
| Neo-late | SUVR | 0.619 (0.571–0.660) | −1.903 (−2.181–−1.654) | 2.673 (2.445–2.931) |

CCC = Lin’s concordance correlation coefficient. Deming regression assumes equal measurement-error variance. Confidence intervals were obtained by participant-level bootstrap resampling. TSE = tau spatial extent; SUVR = standardized uptake value ratio; LoA = limts of agreement; CI = confidence interval; MTL = medial temporal lobe.

#### **Supplemental Table 23. Comparison of nRMSE between TSE and SUVR.**

| Region | SUVR nRMSE, % (95% CI) | TSE nRMSE, % (95% CI) | Ratio (TSE/SUVR) | P value |
| --- | --- | --- | --- | --- |
| MTL | 10.78 (9.81–11.86) | 21.75 (20.23–23.28) | 2.02 | <0.001 |
| Neo-late | 6.96 (5.69–8.16) | 11.74 (10.75–12.76) | 1.69 | <0.001 |

Values are presented as nRMSE (95% confidence interval). P values from Wilcoxon signed-rank test. nRMSE = normalized root mean square error; TSE = tau spatial extent; SUVR = standardized uptake value ratio; CI = confidence interval; MTL = medial temporal lobe.
